# The SCCIP-N Trial Protocol: A Randomized Controlled Trial of a Nurse-Led Family Intervention with Hybrid Psychosocial and Physiological Outcome Assessment in Pediatric Cancer

**DOI:** 10.64898/2025.12.30.25343247

**Authors:** Xiaorou Zeng, Ying Kong, Ya Shi, Ying Ye, Can Gu

**Affiliations:** Xiangya School of Nursing, Central South University, Changsha, China; School of Nursing, Xinjang Medical University, Urumqi, China

**Keywords:** Pediatric Cancer, Post-traumatic stress symptoms, Nurse-led intervention, Randomized controlled trial, Family-based intervention

## Abstract

**Background:** Post-traumatic stress symptoms (PTSS) are prevalent among children with cancer and their families. Although family-based interventions such as the Surviving Cancer Competently Intervention Program (SCCIP) are effective, their reliance on mental health specialists limits scalability, highlighting the need for nurse-led approaches.

**Methods and analysis:** This multicenter randomized controlled trial will enroll 110 families of children with cancer to evaluate the efficacy of a nurse-led adaptation, the SCCIP-N. Participants from Hunan and Xinjiang will be randomly assigned (1:1) to the intervention or an active control group. The study employs a hybrid outcome assessment strategy, integrating validated psychosocial measures with objective physiological monitoring. Assessments occur at baseline (T0), 1 week (T1), 4 weeks (T2), and 8 weeks (T3) post-intervention. The primary outcome is PTSS in children and caregivers. Secondary outcomes include posttraumatic growth, cognitions, anxiety, depression, family management, resilience, quality of life, and sleep quality. Objective physiological data (e.g., sleep, heart rate) from primary caregivers will be continuously collected using the Huawei Band 9. A process evaluation will follow the RE-AIM framework. Data will be analyzed according to intention-to-treat principles using linear mixed-effects models.

**Discussion:** This trial will evaluate the efficacy of a novel, nurse-led family intervention (SCCIP-N) for pediatric oncology families. By combining psychosocial and physiological assessment, it aims to provide a comprehensive, multidimensional evaluation of intervention effects. If effective, SCCIP-N could be integrated into routine care to improve family psychological outcomes and inform future implementation strategies.

**Trial registration:** ChiCTR, ChiCTR2500114338. Registered on Dec 10, 2025.

## Background

Childhood cancer has emerged as a critical public health issue worldwide, posing a severe threat to children’s health [1]. According to statistics from the International Agency for Research on Cancer (IARC), approximately 200,000 children were diagnosed with malignant solid tumors globally in 2022, with leukemia being the most common (31.9%), followed by brain and central nervous system tumors (12.2%) and non-Hodgkin lymphoma (8.7%) [2]. As the world’s most populous developing country, China bears a particularly heavy burden, with approximately 40,380 new cases annually among children and adolescents aged 0–19, ranking second globally [3]. Moreover, the Lancet Oncology Commission predicts that over the next 30 years, about 13.7 million children worldwide will be diagnosed with malignant tumors, with cumulative deaths potentially reaching 11.1 million [4].

Beyond the serious threat to physical health, Pediatric Cancer exerts profound psychosocial impacts on patients and their families [5]. Studies indicate that children frequently experience negative emotions such as anxiety, depression, and fear, while their parents report high levels of traumatic reactions, collectively impairing family quality of life [6, 7]. These reactions often cluster as post-traumatic stress symptoms (PTSS), which include intrusive memories, avoidance behaviors, and hyperarousal in response to the cancer experience. Cancer-related PTSS are prevalent, affecting approximately 44% of children and up to 68% of parents, meaning nearly every family experiences this distress [8, 9]. Persistent PTSS can compromise children’s social functioning and treatment adherence, exacerbate parental anxiety and depression, and significantly diminish overall family well-being [10], a pattern also observed in Chinese contexts [11].

To address cancer-related PTSS, cognitive-behavioral therapy (CBT) has been recommended as a first-line trauma intervention strategy in international guidelines [12]. Evidence underscores its efficacy for both children with cancer and their parents. For children, CBT-based interventions have been shown to improve mental health and quality of life, with benefits extending to long-term survivors [13]. A systematic review confirms CBT’s significant role in alleviating a range of distress symptoms in pediatric populations, including PTSS, pain, anxiety, and depression [14]. For parents, structured interventions such as Cognitive-Behavioral Stress Management (CBSM) can effectively reduce anxiety, depression, and post-traumatic stress [15–17]. Furthermore, the adaptable principles of CBT provide a valuable framework for supporting caregivers’ psychosocial adaptation to their challenging roles [15–17]. The Surviving Cancer Competently Intervention Program (SCCIP), developed by Kazak’s team, integrates cognitive-behavioral and family therapy techniques to reduce PTSS and anxiety while improving family functioning [18]. An RCT with 136 U.S. families demonstrated its efficacy in reducing distress, improving family management, and confirming its feasibility and acceptability [6].

Our team previously culturally adapted SCCIP for Chinese families. While acceptable, its feasibility was limited by a reliance on specialized mental health providers, hindering scalability within routine nursing care. This highlights the need for a nurse-led delivery model. Nurses, by virtue of their continuous contact with families, possess unique advantages in providing psychosocial support [19]. Although nurse-led interventions have shown benefits in adult oncology [20, 21], in improving resilience and satisfaction in pediatric settings [22, 23], and in enhancing treatment adherence [24], rigorous evidence for nurse-led, family-based psychosocial interventions targeting PTSS in pediatric cancer remains scarce. Critically, existing studies predominantly rely on self-reported psychological outcomes. A significant gap is the lack of integrated, objective physiological measures to complement psychosocial assessments and provide a more comprehensive understanding of intervention effects. Leveraging nurses’ pivotal role and addressing this methodological gap, we developed a nurse-led adaptation of SCCIP (SCCIP-N).

Therefore, this study will conduct a multicenter randomized controlled trial to evaluate the SCCIP-N intervention. Crucially, and as a key methodological advancement, the trial will employ a hybrid psychosocial and physiological outcome assessment strategy. This approach combines validated self-report measures with objective, continuous physiological data (e.g., sleep, heart rate) collected via wearable devices from primary caregivers. We aim to determine the intervention’s effects on PTSS and related psychosocial outcomes while exploring its impact on physiological stress correlates, thereby offering a multidimensional evaluation of efficacy in Chinese pediatric oncology families.

## Methods and analysis

### Trial Design

This project is a two-arm parallel randomized controlled trial designed to evaluate the effectiveness of the SCCIP-N in reducing PTSS among families of children with cancer. Following informed consent, eligible families will be randomly assigned in a 1:1 ratio to either the intervention group or the control group. The brief intervention consists of four sessions delivered within a single day. The primary outcome and the secondary outcomes will be surveyed at four time points: at baseline (T0), at 1 week post-intervention (T1), at 4-week follow-ups (T2) and 8-week follow-ups (T3). Fig1 presents the schedule of participant enrolment, interventions, and assessments. The study flowchart is shown in Fig2. The study protocol has been developed and reported in accordance with the Standard Protocol Items: Recommendations for Interventional Trials (SPIRIT) 2025 guidelines [25] (S1 File).

**Fig 1.**
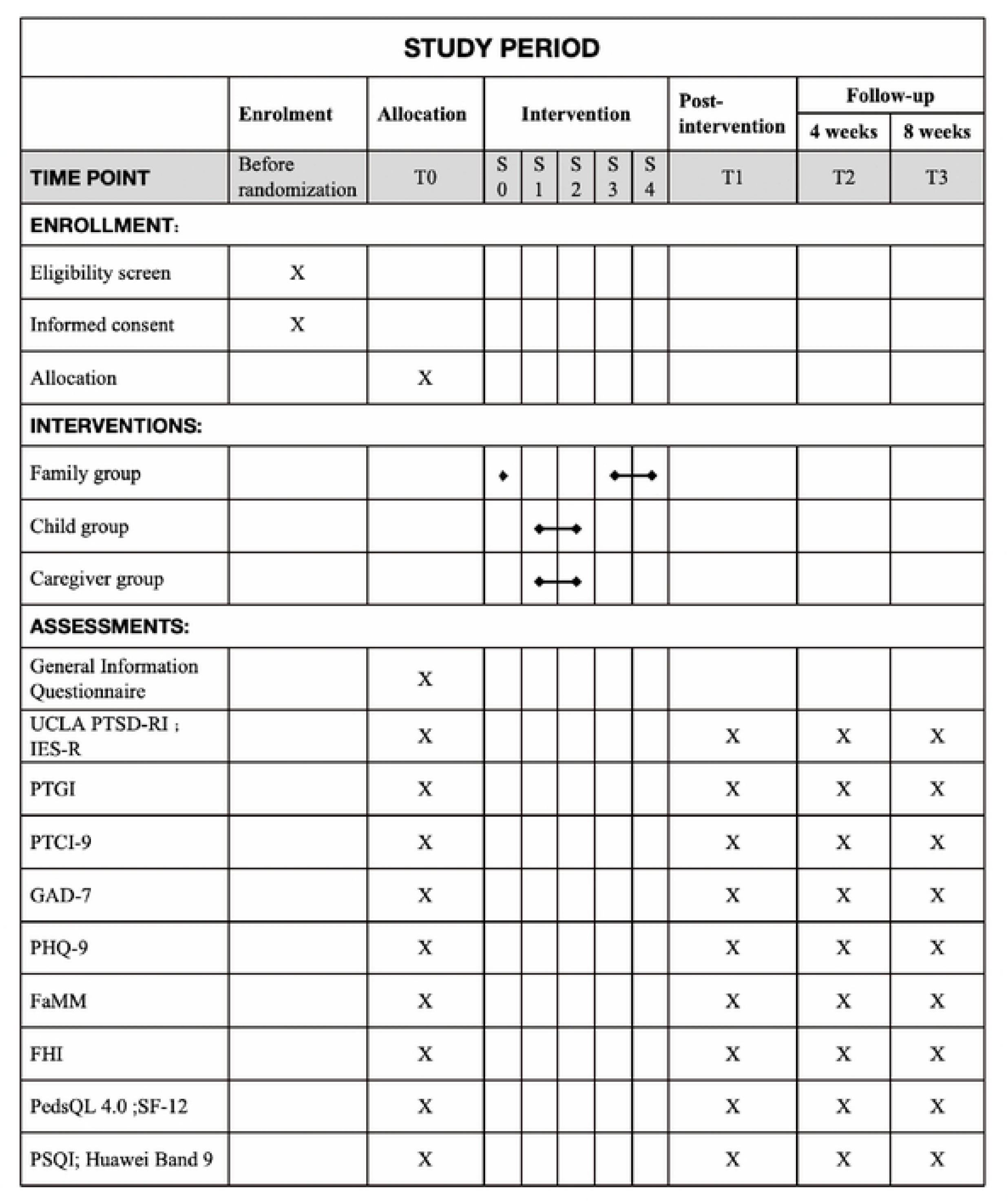
SPIRIT schedule of participant enrolment, interventions, and assessments. Abbreviations: UCLA PTSD-RI, University of California at Los Angeles Posttraumatic Stress Disorder Reaction Index for DSM-IV, Child Version; IES-R, Impact of Event Scale–Revised; PTGI, Posttraumatic Growth Inventory; PTCI-9, Brief Version of the Posttraumatic Cognitions Inventory; GAD-7, 7-item Generalized Anxiety Disorder scale; PHQ-9, Patient Health Questionnaire–9; FaMM, Family Management Measure; FHI, Family Hardiness Index; PedsQL 4.0, Pediatric Quality of Life Inventory Generic Core Scales; SF-12, 12-Item Short Form Health Survey; PSQI, Pittsburgh Sleep Quality Index.

**Fig 2.**
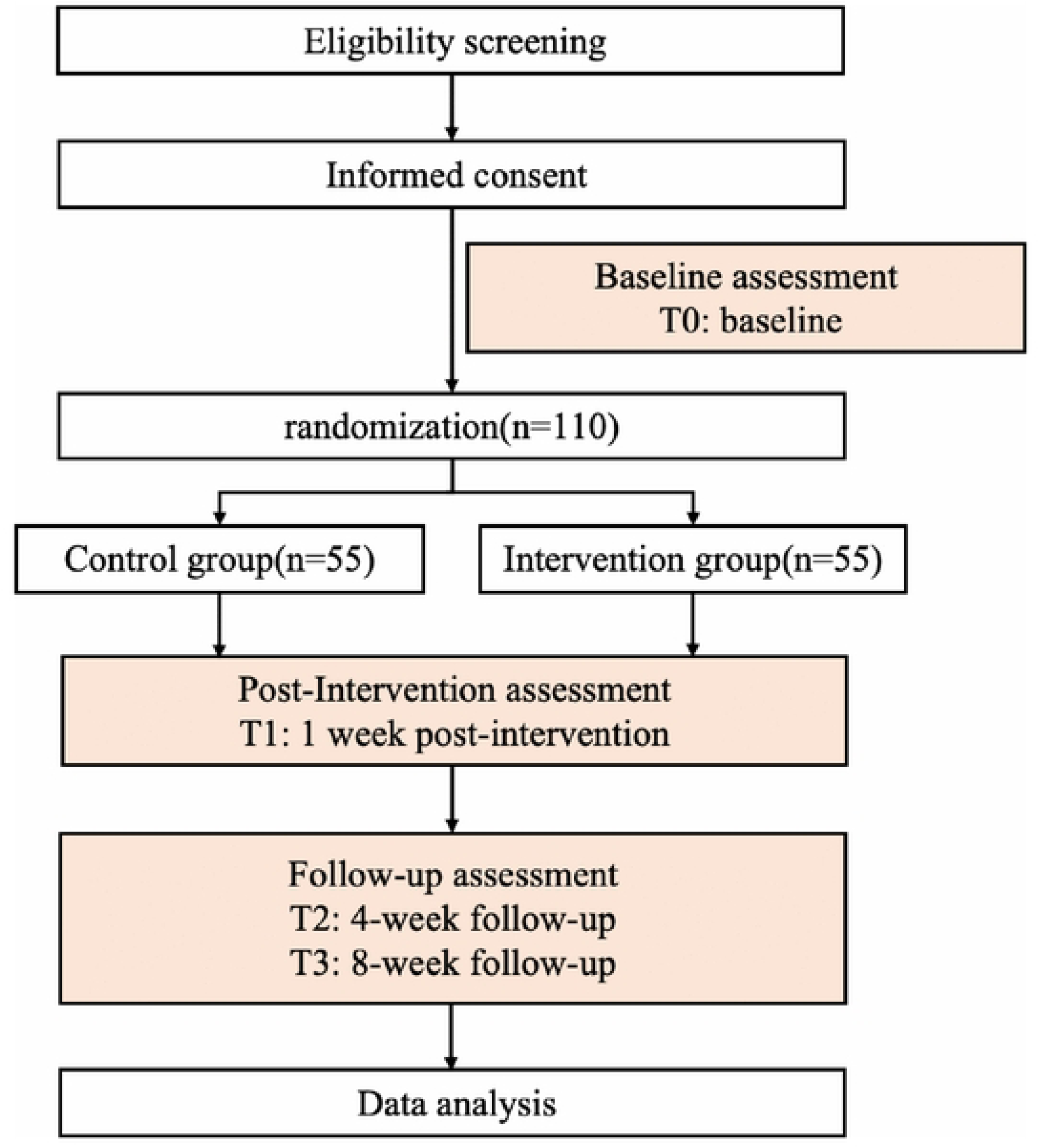
Flowchart of study design.

### Study setting

The trial will be undertaken in two regions of China — Hunan Province in Central China and the Xinjiang Uygur Autonomous Region in Northwest China, with all interventions conducted in the inpatient departments of several tertiary A hospitals.

### Sample size estimation

The sample size is estimated based on changes in children’s PTSS measured by the PTSD-RI total score in a pilot study. In the pilot, the mean change from baseline to 2 months post-intervention was 5.73 ± 8.19 in the intervention group and 1.07 ± 8.41 in the control group, corresponding to a mean difference of 4.66 [26]. The pooled standard deviation of the change scores was 8.28, yielding an effect size of Cohen’s d = 0.56.

Considering analysis using the independent two-sample Student’s t-test, and based on this effect size, the required sample size is calculated using G*Power 3.1 software. The analysis indicates that 44 participants per group are needed (α = 0.05 and power = 0.80). To account for an estimated 20% attrition rate, the final adjusted sample size is set at 55 participants per group, resulting in a total of 110 participants.

### Participants

To be eligible for this study, families of children with cancer (including the child and at least one primary caregiver) must meet the following criteria:

#### Inclusion criteria

1. Child diagnosed with a malignant tumor based on imaging, pathological, or cytological examinations, in accordance with the International Classification of Childhood Cancer, Third Edition [27].
2. Child aged 7–18 years and currently receiving active cancer treatment (e.g., chemotherapy, radiotherapy, or targeted therapy).
3. Primary caregiver identified as a family member with a biological or legal relationship to the child and serving as the main caregiver during hospitalization.
4. Child and primary caregiver have adequate cognitive and communication abilities.
5. Child and primary caregiver voluntarily agree to participate and provide written informed consent.

#### Exclusion criteria

1. Child with comorbid severe physical conditions
2. Child or primary caregiver has severe psychiatric or cognitive disorders preventing participation in the study.
3. Child or primary caregiver has participated in similar psychological or family intervention programs within the past 6 months.

### Recruitment

Participant recruitment is scheduled to occur from December 2025 to June 2026. Multiple complementary strategies will be employed to identify and engage eligible participants. With approval from the relevant hospital departments, study roll-up banners will be placed in public areas designated by the participating departments, and SCCIP-N flyers will be distributed to families who may be interested in or eligible for the study. Potentially eligible families will also be identified through medical record screening and direct referrals from healthcare providers who are familiar with the inclusion criteria. To further enhance recruitment, word-of-mouth and snowball sampling methods will be applied. Families already enrolled in the study may, on a voluntary basis, refer other potentially eligible families to participate.

### Randomisation and blinding

Eligible families will be randomized at the family level to either the intervention or control group with a 1:1 allocation ratio after providing informed consent and completing baseline assessments. The randomization sequence is generated in advance by an independent statistician using a computer program with randomly varying block sizes, which helps reduce predictability. Each sequence is consecutively numbered and securely held by an independent coordinator who has no role in recruitment, intervention delivery, or outcome assessment.

Once a family completes consent and baseline assessment, they are assigned to the group corresponding to their enrolment number in the pre-generated sequence. Recruitment staff and intervention providers remain blinded until this point, ensuring allocation concealment. Randomization proceeds in batches until the target sample size is reached.

Blinding of participants and intervention providers is not feasible due to the obvious differences between the study conditions. Nevertheless, outcome assessors remain blinded throughout the study to minimize assessment bias.

### Intervention

#### Intervention group

The cultural adaptation of the SCCIP-N for Chinese pediatric oncology families was completed in a preliminary phase of the study. Based on qualitative interviews with families and healthcare providers, expert consultation, and pilot testing, the research team conducted a standardized translation and localization of the original manual to ensure cultural relevance and feasibility. The final Chinese version retained the core theoretical foundations of the intervention, including cognitive–behavioral and family systems approach.

The SCCIP-N is a structured, brief intervention delivered in a single day, lasting approximately 90-120 minutes, and designed for small groups of 2 to 5 families. The program includes one orientation session and four structured sessions, with full details available in Table 1.

**Table 1.** Session Structure and Content Overview: The SCCIP-N intervention.

| Session | Theme | Content | Group |
| --- | --- | --- | --- |
| Orientation | Introduction and Program Overview” | 1、 Welcome & Team Introductions<br>2、 Overview of Activities and Format<br>3、 Group Allocation & Venue Guidance | Family group |
| Session1 | How cancer has affected me and my family | 1、 Ice-breaker activity<br>2、 Understanding Post-traumatic stress<br>3、 Identifying Unhappy Memories:<br>Select three cancer-related distressing memories using team-developed cards.<br>Discuss how these memories have affected themselves. | Child group<br>Primary caregiver group |
| Session2 | Coping skills. | 1、 The Adversity-Beliefs-Consequences (ABC) model<br>2、 Reframing Negative Beliefs<br>3、 Practice with examples (e.g., chemotherapy, healthcare staff rounds)<br>4、 Application of ABC model and Reframing to unhappy memories | Child group<br>Primary caregiver group |
| Session3 | Getting on with life—Cancer, adolescents, and families. | 1、 Review of previous sessions and comic introduction<br>2、 General group discussion: How has the illness affected the entire family?<br>3、 Multiple family discussion groups (MFDGs):<br>Caregivers: Have your thoughts about the illness affected your family life?<br>Children: What cancer-related matters are truly bothering you? | Family group |
| Session4 | Pulling it all together—Family health and our future. | 1、 Discussion on cancer's life occupation and facilitation of family recognition, acceptance, and contextualization<br>2、 Developing and sharing family future plans<br>3、 Activity feedback | Family group |

In the first two sessions, children and their primary caregivers participate in separate child and caregiver groups, with content tailored to the specific needs of each population. While the core components remain consistent, child sessions are adapted using age-appropriate materials such as cartoons, stories, and role-play. The final two sessions involve family group activities, in which children and their primary caregivers participate together. These sessions aim to enhance communication and promote shared coping among family members.

After the intervention, facilitators will establish a WeChat group for each cohort. These groups provide ongoing psychological support during the follow-up phase, facilitate experience sharing, and offer timely feedback, thereby extending the benefits of the intervention and strengthening both peer and family interactions.

The intervention will be delivered by two trained registered nurses, with support from two assistants. Prior to implementation, all facilitators will complete a 4-hour training program covering the theoretical principles of nurse-led psychological and family interventions, session procedures, and safety monitoring. Each facilitator must observe at least one complete intervention session before conducting sessions independently. This step ensures both fidelity to the intervention protocol and the quality of delivery.

#### Control group

Participants in the control group receive routine psychological support alongside standard medical care. This support includes emotional assistance through daily interactions and active listening with children and their caregivers; provision of health education related to disease treatment and nursing (e.g., dietary recommendations, lifestyle guidance, and infection and bleeding prevention). Upon completion of the study, families expressing interest in SCCIP-N will be offered the intervention.

### Outcome Measures

#### Multimodal Outcome Assessment Strategy

To comprehensively evaluate the intervention’s impact, this trial employs a hybrid assessment strategy that combines well-validated patient-reported outcome measures (PROMs) with objective, continuous physiological monitoring. This approach allows for a more robust and multidimensional understanding of changes in post-traumatic stress and family adaptation. A summary of all outcome variables and their corresponding measurement instruments for both children and primary caregivers is presented in Table 2.

**Table 2.** Summary of Outcome Measures for Children and Caregivers.

| Outcome | Measure | Respondent |  |
| --- | --- | --- | --- |
|  |  | Child | Caregiver |
| Sociodemographic and Clinical Variables | General Information Questionnaire |  | X |
| Posttraumatic stress symptoms* | UCLA PTSD-RI | X |  |
|  | IES-R |  | X |
| Posttraumatic growth | PTGI | X | X |
| Posttraumatic cognitions | PTCI-9 | X | X |
| Anxiety | GAD-7 | X | X |
| Depression | PHQ-9 | X | X |
| Family management | FaMM |  | X |
| Family resilience | FHI |  | X |
| Health-related quality of life | PedsQL 4.0 | X |  |
|  | SF-12 |  | X |
| Sleep and physiological indicators | PSQI | X |  |
|  | Huawei Band 9 |  | X |

Note: Note: The measures marked with an asterisk (\*) constitute the primary outcome. UCLA PTSD-RI, The University of California at Los Angeles Posttraumatic Stress Disorder Reaction Index for DSM-IV, Child Version; IES-R, Impact of Event Scale–Revised; PTGI, Posttraumatic Growth Inventory; PTCI-9, The Brief Version of the Posttraumatic Cognitions Inventory; GAD-7, 7-item Generalized Anxiety Disorder scale; PHQ-9, Patient Health Questionnaire–9; FaMM, Family Management Measure; FHI,
Family Hardiness Index; PedsQL 4.0, PedsQL 4.0 Generic Core Scales; SF-12, 12-Item Short Form Health Survey; PSQI, Pittsburgh Sleep Quality Index.

#### Primary Outcome

The primary outcome of this trial is PTSS in both children and their primary caregivers.

##### PTSS in Children

The University of California at Los Angeles Posttraumatic Stress Disorder Reaction Index for DSM-IV, Revision 1, Child Version (UCLA PTSD-RI)[28] was utilized to assess PTSS in children. The scoring is based on the diagnostic criteria outlined in the Diagnostic and Statistical Manual of Mental Disorders, Fourth Edition (DSM-IV). The UCLA PTSD-RI is regarded as the current gold standard for evaluating PTSD symptoms in pediatric populations[29]. The scale consists of 20 items that encompass three symptom clusters: intrusion, avoidance, and hyperarousal. Each item is rated on a 5-point scale, ranging from “none of the time”(0) to “most of the time” (4), with higher scores indicating more severe PTSS. In this study, the Chinese version translated and validated by Fu et al.[30] will be employed. This version has demonstrated robust reliability and validity.

##### PTSS in Primary Caregivers

The Impact of Event Scale–Revised (IES-R) is a validated self-report instrument for assessing post-traumatic stress symptoms[31]. The questionnaire comprises 22 items that address three symptom clusters: intrusion, avoidance, and hyperarousal. Each item is rated on a 5-point scale (0 = not at all; 1 = a little bit; 2 = moderately; 3 = quite a bit; 4 = extremely), with total scores ranging from 0 to 88. Higher scores indicate more severe PTSS, and a total IES-R score of ≥33 suggests probable PTSD[32]. In this study, we will utilize the Chinese version of the CIES-R, which has exhibited strong psychometric properties[32].

#### Secondary Outcomes

##### Posttraumatic Growth

The Posttraumatic Growth Inventory (PTGI)[32] is a self-assessment tool designed to measure posttraumatic growth (PTG) and has been widely utilized among cancer children[33] and their caregivers[34]. In this study, the Chinese version of the PTGI (C-PTGI) will be employed. The C-PTGI assesses five key dimensions: relationships with others, new possibilities, personal strength, spiritual growth, and appreciation of life. Each of the 20 items is rated on a scale from 0 (not at all) to 5 (very much), effectively capturing the individual’s growth. The total score ranges from 0 to 100, with higher scores indicating greater PTG. The scale demonstrates strong psychometric properties, with a reported Cronbach’s α coefficient of 0.874 for the total scale[35].

##### Posttraumatic Cognitions

The Brief Version of the Posttraumatic Cognitions Inventory (PTCI-9) is a reliable and valid self-report questionnaire developed to assess posttraumatic cognitions. It was developed by Wells et al.[36] as a modification of the original PTCI[37].The scale includes nine items covering three main domains: Negative Cognitions about the Self, Negative Cognitions about the World, and Self-Blame. Each item is rated on a 7-point Likert scale ranging from 1 (totally disagree) to 7 (totally agree), resulting in a total score between 9 and 63, where higher scores indicate more severe posttraumatic negative cognitions. The PTCI-9 has been validated in Chinese adolescent and adult populations and exhibits acceptable internal consistency[38].

##### Anxiety and Depression

The 7-item Generalized Anxiety Disorder scale (GAD-7)[39] is a brief screening tool used to identify anxiety and assess the severity of anxiety symptoms. Each item is rated on a 4-point scale: 0 (never), 1 (several days), 2 (more than half the days), and 3 (nearly every day), for a total score of 0 to 21. A higher score indicates more severe anxiety symptoms.

The Patient Health Questionnaire-9 (PHQ-9)[40] is a widely employed instrument for evaluating depressive symptoms. It consists of nine items, with responses rated on a 4-point Likert scale based on the frequency of symptoms experienced in the past two weeks (0 = “not at all” to 3 = “nearly every day”). Total scores range from 0 to 27, with higher scores signifying greater symptom severity.

##### Family Management

The Family Management Measure (FaMM)[41] assesses how families manage the care of a child with a chronic condition and the impact of this condition on family life. The scale contains 53 items across six dimensions: Child’s Daily Life, Condition Management Ability, Condition Management Effort, Family Life Difficulty, View of Condition Impact, and Parental Mutuality (43 items for single-parent families, excluding the Parental Mutuality subscale). Items are rated on a 5-point Likert scale (1 = “strongly disagree” to 5 = “strongly agree”). The scale was translated into Chinese by Zhang et al.[41] and validated among Chinese families with children facing chronic conditions, demonstrating strong psychometric properties.

##### Family Resilience

The Family Hardiness Index (FHI)[42] consists of 20 items assessing three dimensions: commitment, challenge, and control. Each item is rated on a 4-point Likert scale ranging from 0 (“completely wrong”) to 3 (“completely correct”), with higher scores indicating greater family hardiness. In this study, the Chinese version of the FHI, translated and validated by Liu et al[43]. will be used. The scale has demonstrated good reliability, with a Cronbach’s α of 0.803, as well as acceptable overall psychometric properties.

##### Health-Related Quality of Life (HRQOL)

The 23-item PedsQL 4.0 Generic Core Scales[44] will be employed to assess the HRQOL of children and adolescents across four dimensions: physical functioning, emotional functioning, social functioning, and school functioning. Each item is rated on a 5-point Likert scale ranging from 0 (“never”) to 4 (“almost always”). Items are reverse-scored and linearly transformed to a 0-100 scale, with higher scores reflecting better HRQOL. The Chinese version of the PedsQL 4.0 has demonstrated satisfactory reliability and validity among pediatric cancer patients[45].

The 12-item Short-Form Health Survey (SF-12)[45] will be utilized to assess the quality of life of primary caregivers of pediatric cancer patients. The SF-12 includes 12 items that encompass eight subscales: general health, physical functioning, bodily pain, role-physical, mental health, vitality, social functioning, and role-emotional. According to the scoring manual, these eight subscales are aggregated into two composite dimensions: the Physical Component Summary (PCS) and the Mental Component Summary (MCS). All scores were transformed to a 0-100 scale, with higher scores indicating better quality of life.

##### Sleep and physiological indicators

The Pittsburgh Sleep Quality Index (PSQI)[46] is a 19-item self-report measure used to assess sleep quality in children with cancer. It covers seven components, with total scores ranging from 0 to 21; higher scores indicate poorer sleep quality. The Chinese version has demonstrated good reliability and validity in pediatric oncology populations [47].

The Huawei Band 9 will be used to assess objective physiological metrics in primary caregivers of children with cancer. Key parameters include nocturnal total sleep time [48], sleep score, mean heart rate, average blood oxygen saturation, and daytime stress levels. These parameters were selected as validated proxies for autonomic nervous system activity and stress-related physiological burden, which are known correlates of psychosocial distress in caregivers. The use of a consumer-grade wearable balances rigorous data collection with feasibility and minimal participant burden in a clinical setting. The device continuously collects data using a photoplethysmography (PPG) sensor [49] and a 9-axis inertial measurement unit (IMU). Relevant parameters are automatically calculated through an integrated multimodal data fusion algorithm. Data is exported via the Huawei Health app for statistical analysis. This approach enables continuous and objective monitoring of significant physiological changes during the intervention without imposing additional burden on participants.

### Data Collection and Management

#### Sociodemographic and Clinical Variables

At the baseline stage, a self-designed general information questionnaire will be used to collect sociodemographic and clinical characteristic data of the study participants. The main contents include: Sociodemographic characteristics of the primary caregiver: age, gender, education level, marital status, occupation, and family type; Clinical characteristics of the child: age, gender, school grade, diagnosis, disease stage/risk level, treatment phase, presence of metastasis, and type of treatment.

#### Process evaluation

The process evaluation will be prespecified by the use of the RE-AIM frameworks[50]. Qualitative and quantitative data will be collected to inform the Reach, Effectiveness, Adoption, Implementation, Maintenance (RE-AIM) of the intervention. Reach of the intervention will be expressed as the absolute and relative number of participating families of children with cancer, together with their demographic and clinical characteristics (described above) to assess representativeness. Effectiveness refers to the impact of the intervention on outcomes such as post-traumatic stress symptoms. Adoption will be assessed through interviews exploring nurses’ willingness and perceived feasibility of applying the intervention in their future clinical practice. Implementation will be evaluated in terms of fidelity and acceptability. Fidelity refers to the degree of consistency in delivering the intervention components according to the study protocol. Acceptability will be assessed using a 5-point Likert satisfaction scale (1 = very dissatisfied, 5 = very satisfied), covering aspects such as the intervention content, format, duration, and facilitator performance, followed by brief focus group interviews about participants’ overall experience. Maintenance, referring to the long-term sustainability of intervention effects, is beyond the scope of this study[51].

#### Data management

All questionnaire data will be recorded in standardized case report forms (CRFs) and collected by trained research staff according to established procedures. Objective physiological indicators will be automatically collected using the Huawei Smart Band. After data collection, two independent data entry personnel will conduct double entry and cross-verification. The electronic data capture system will include real-time validation features to automatically identify missing or inconsistent values. To minimize data loss, regular communication and flexible follow-up scheduling will be implemented. Paper CRFs will be securely stored in locked cabinets, while de-identified (coded) electronic data will be stored on a locally encrypted server to ensure data security and confidentiality. Access to the complete dataset will be restricted to authorized research team members only. All data will be retained for at least three years after the study is completed.

### Statistical analysis

Baseline sociodemographic and clinical characteristics will be summarized as means ± SD for continuous variables that are normally distributed, or as medians (IQR) for continuous variables that are not normally distributed. Between-group comparisons at baseline will be conducted using independent samples t-tests for normally distributed continuous variables, Mann-Whitney U tests for non-normally distributed continuous variables, χ²tests for categorical variables with sufficient expected counts, and Fisher’ s exact tests for categorical variables with small expected counts.

The primary outcome, PTSS scores of children and their primary caregivers, will be analyzed using linear mixed-effects models (LMM) for continuous outcomes over time (T0, T1, T2, T3). Fixed effects include group (intervention vs. control), time, and group×time interaction, with participant-level intercepts as random effects to account for within-subject correlations. For categorical outcomes (e.g., reaching clinical cutoff), generalized linear mixed-effects models (GLMM) with random intercepts will be used. Effect sizes (Cohen’s d) will be calculated to quantify between-group differences at each time point. Secondary outcomes, including anxiety, depression, quality of life, and family functioning scores, will be analyzed using the same mixed-effects models as described for the primary outcome. To explore the relationship between psychosocial changes and physiological metrics, correlation analyses (e.g., Pearson’s or Spearman’s) will be conducted between key change scores (e.g., ΔIES-R, ΔPHQ-9) and changes in physiological parameters (e.g., mean sleep score, nighttime heart rate) across assessment time points.

In addition, missing data are almost inevitable. For a small proportion of missing values, conditional mean imputation will be applied. When missingness is substantial, multiple imputation will be performed. All statistical tests will be two-sided with a significance level of p < 0.05, and 95% confidence intervals (CI) will be reported. Data processing and analysis will be carried out using the statistical package SPSS, V.26.0.

### Data and Safety Monitoring

Given that this trial evaluates a low-risk, nurse-led psychosocial and supportive care intervention that does not involve investigational drugs, medical devices, or procedures altering physiological processes, an independent Data Monitoring Committee (DMC) is not required. The intervention represents an extension of standard nursing care and is anticipated to pose minimal risk to participants, defined as risks no greater than those ordinarily encountered in daily life or during routine psychological or supportive care. In view of the low-risk and supportive nature of the intervention, no formal interim analyses for efficacy or futility are planned.

#### Patient and public involvement

Prior to the trial, primary caregivers of children with cancer were consulted through qualitative interviews to inform the cultural adaptation and design of the nurse-led SCCIP intervention, including its content, format, and delivery. Insights from these interviews enhanced the feasibility and acceptability of the intervention for Chinese pediatric oncology families. Patients or the public were not involved in the conduct, analysis, or reporting of the trial.

### Ethics and dissemination

This study was approved by the Ethics Review Committee of Xiangya School of Nursing, Central South University (Approval Number: E2025172) and the Medical Ethics Committee of Central South University (Approval Number: CSUMEC-E2025011).

All participants will receive detailed oral and written information about the study and will sign an Informed Consent Form prior to participation. Participation is voluntary, and participants may withdraw from the study at any time without penalty or loss of benefits to which they are otherwise entitled. The study findings will be disseminated through publications in peer-reviewed journals, presentations at academic conferences, and, where appropriate, communication through media channels to enhance public awareness.

### Protocol Amendments

Any substantial modifications to the study protocol that may impact the conduct of the trial, participant safety, study objectives, or outcomes will be reviewed and approved by the relevant ethics committee prior to implementation. All approved protocol amendments will be communicated to relevant stakeholders, including investigators, trial staff, and, where applicable, trial registries. Participants will be informed of protocol changes when necessary, in accordance with ethical and institutional requirements.

#### Study status

This study is registered in the Chinese Clinical Trial Registry (ChiCTR, Registration NO. ChiCTR2500114338). Recruitment of participants began on December 20, 2025, and is expected to be completed by June 20, 2026. Data collection is anticipated to be completed by December 20, 2026.

## Discussion

This study employs a prospective randomized controlled trial to evaluate the effectiveness of the SCCIP-N intervention for families of children with cancer. The intervention is guided by a standardized manual that has been culturally adapted and pilot-tested for the Chinese context, balancing methodological rigor with local relevance. To our knowledge, this is among the first randomized controlled trials in pediatric oncology to combine a nurse-led delivery model with a family-based psychosocial approach [52]. Leveraging the expertise of registered nurses, the model addresses the shortage of mental health professionals while providing accessible, compassionate psychosocial support within oncology care. The family-based component treats the family as an interconnected system, ensuring that both children with cancer and their primary caregivers receive comprehensive support, and the group format promotes communication and mutual support among multiple families[53]. The study evaluates a range of multidimensional outcomes, including subjective measures (posttraumatic stress symptoms, emotional well-being, family functioning, and quality of life) and objective physiological indicators (e.g., sleep parameters), providing a comprehensive assessment of the intervention’s impact at both individual and family levels.

This investigation is not without its limitations. The single-blinded design may introduce performance and expectation biases. The single-day intervention format, while feasible, may restrict the depth of learning and the sustainability of outcomes compared with longer or repeated protocols [54]. Furthermore, most commercially available wearable devices are designed for adults ( ≥ 18 years), which limits the collection of physiological data from pediatric participants. This limits our ability to capture the direct physiological impact of the intervention on the child patients.

If effective, SCCIP-N could be recommended as a low-risk, easily implementable intervention to reduce posttraumatic stress symptoms in families of children with cancer and integrated into routine oncology care. Future studies may explore extending the duration or frequency of the intervention to enhance learning and maintain effects. Incorporating mobile health platforms could provide continuous psychosocial support, particularly for fathers, siblings, and families living in remote areas who have limited access to hospital-based care[55].

## Data Availability

No datasets were generated or analysed during the current study. All relevant data from this study will be made available upon study completion.

## Acknowledgments

We thank the participating hospitals and the research personnel involved in the design and preparation of this study for their support and cooperation.

## Supporting information

### S1 File. SPIRIT 2025 editable checklist

### S2 File. Ethical approval letter

## Notes

**Funding sources:** This work was supported by the National Natural Science Foundation of China (Grant No. 82272924 to Can Gu).

### Competing Interest Statement

The authors have declared no competing interest.

### Clinical Trial

ChiCTR2500114338

### Clinical Protocols

https://www.chictr.org.cn/

### Funding Statement

Yes

## References

1. Ortiz R, Vásquez L, Giri B, Kapambwe S, Dille I, Mahmoud L, et al. Developing and sustaining high-quality care for children with cancer: the WHO Global Initiative for Childhood Cancer. Revista Panamericana De Salud Publica-Pan American Journal of Public Health. 2023;47. doi: 10.26633/rpsp.2023.164. PubMed PMID: WOS:001129035400001.

2. Peng YY, Xu L, Gu C, Ma GY, Zhang ZT, Zhang YL, et al. Prevalence and associated factors of post-traumatic stress symptoms in hospitalised children with cancer and their parents in South China: A multicentred cross-sectional study. Asia-Pacific Journal of Oncology Nursing. 2024;11(10). doi: 10.1016/j.apjon.2024.100568. PubMed PMID: WOS:001336985000001.

3. Ni X, Li Z, Li XP, Zhang X, Bai GL, Liu YY, et al. Socioeconomic inequalities in cancer incidence and access to health services among children and adolescents in China: a cross-sectional study. Lancet. 2022;400(10357):1020–32. doi: 10.1016/s0140-6736(22)01541-0. PubMed PMID: WOS:000875091000003.

4. Sullivan CE, Challinor J, Pergert P, Afungchwi GM, Downing J, Morrissey L, et al. Strengthening the global nursing workforce for childhood cancer. Lancet Oncology. 2020;21(12):1550–2. doi: 10.1016/s1470-2045(20)30425-3. PubMed PMID: WOS:000599899200032.

5. Ricart B, Carter JS. Commentary: Increasing generalizability of parent psychosocial functioning within the context of pediatric chronic pain. Journal of Pediatric Psychology. 2024;49(5):318–20. doi: 10.1093/jpepsy/jsae019. PubMed PMID: WOS:001188301800001.

6. Kazak AE, Alderfer MA, Streisand R, Simms S, Rourke MT, Barakat LP, et al. Treatment of posttraumatic stress symptoms in adolescent survivors of childhood cancer and their families: A randomized clinical trial. Journal of Family Psychology. 2004;18(3):493–504. doi: 10.1037/0893-3200.18.3.493. PubMed PMID: WOS:000223792900010.

7. Landolt MA, Ystrom E, Sennhauser FH, Gnehm HE, Vollrath ME. The mutual prospective influence of child and parental post-traumatic stress symptoms in pediatric patients. Journal of Child Psychology and Psychiatry. 2012;53(7):767–74. doi: 10.1111/j.1469-7610.2011.02520.x. PubMed PMID: WOS:000305060800007.

8. Afzal N, Ye SY, Page AC, Trickey D, Lyttle MD, Hiller RM, et al. A systematic literature review of the relationship between parenting responses and child post-traumatic stress symptoms. European Journal of Psychotraumatology. 2023;14(1). doi: 10.1080/20008066.2022.2156053. PubMed PMID: WOS:000899295600001.

9. Allbaugh LJ, George G, Klengel T, Profetto A, Marinack L, O’Malley F, et al. Children of trauma survivors: Influences of parental posttraumatic stress and child-perceived parenting. J Affect Disord. 2024;354:224–31. Epub 20240313. doi: 10.1016/j.jad.2024.03.067. PubMed PMID: 38490588.

10. Ma J, Qian HZ, Peng YY, Xiang YL, Yang MH, Hahne J, et al. Efficacy of a smartphone-based care support programme in improving post-traumatic stress in families with childhood cancer: protocol of a randomised controlled trial. Bmj Open. 2022;12(9). doi: 10.1136/bmjopen-2021-060629. PubMed PMID: WOS:000859948800025.

11. Yang Y, He X, Chen J, Tan X, Meng J, Cai R, et al. Posttraumatic stress symptoms in Chinese children with ongoing cancer treatment and their parents: Are they elevated relative to healthy comparisons? Eur J Cancer Care (Engl). 2022;31(2):e13554. Epub 20220207. doi: 10.1111/ecc.13554. PubMed PMID: 35129840.

12. Xiang LA, Wan HW, Zhu Y. Effects of cognitive behavioral therapy on resilience among adult cancer patients: a systematic review and meta-analysis. Bmc Psychiatry. 2025;25(1). doi: 10.1186/s12888-025-06628-3. PubMed PMID: WOS:001439684100006.

13. Seitz DCM, Knaevelsrud C, Duran G, Waadt S, Loos S, Goldbeck L. Efficacy of an internet-based cognitive-behavioral intervention for long-term survivors of pediatric cancer: a pilot study. Supportive Care in Cancer. 2014;22(8):2075–83. doi: 10.1007/s00520-014-2193-4. PubMed PMID: WOS:000339339300009.

14. Pelcastre-Neri A, García-Solano B, Herrera-Paredes JM, Carreño-Moreno S, González-Rivera G, Ruvalcaba-Ledezma JC. Interventions for psychological problems in caregivers of children with cancer: a systematic review. Acta Paulista De Enfermagem. 2025;38. doi: 10.37689/acta-ape/2025AR001835i. PubMed PMID: WOS:001611612800001.

15. Wang L, Duan H, Zuo HM, Wang ZY, Jiao SL, Liu YL, et al. Cognitive-behavioral stress management relieves anxiety, depression, and post-traumatic stress disorder in parents of pediatric acute myeloid leukemia patients: a randomized, controlled study. Hematology. 2024;29(1). doi: 10.1080/16078454.2023.2293498. PubMed PMID: WOS:001125162300001.

16. Melesse TG, Li WHC, Chau JPC, Yimer MA, Gidey AM, Yitayih S. Cognitive-Behavioral Intervention for Children With Hematological Cancer Receiving Chemotherapy: A Randomized Controlled Trial. Psycho-Oncology. 2025;34(1). doi: 10.1002/pon.70086. PubMed PMID: WOS:001399994100001.

17. Salley CG, McDonnell GA, Parris KR. Applying Principles of Cognitive Behavioral Therapy to Support Caregivers of Children With Cancer. Cognitive and Behavioral Practice. 2024;31(3):413–22. doi: 10.1016/j.cbpra.2024.01.004. PubMed PMID: WOS:001267688000001.

18. Kazak AE, Simms S, Barakat L, Hobbie W, Foley B, Golomb V, et al. Surviving Cancer Competently Intervention Program (SCCIP): A cognitive-behavioral and family therapy intervention for adolescent survivors of childhood cancer and their families. Family Process. 1999;38(2):175–91. PubMed PMID: WOS:000080705700005.

19. Huang HT, Zhang XN, Tu L, Zhang L, Chen H. Effectiveness of nurse-led self-care interventions on quality of life, social support, depression and anxiety among people living with HIV: A systematic review and meta-analysis of randomized controlled trials. International Journal of Nursing Studies. 2025;161. doi: 10.1016/j.ijnurstu.2024.104916. PubMed PMID: WOS:001333039700001.

20. Propp KM, Apker J, Ford WSZ, Wallace N, Serbenski M, Hofmeister N. Meeting the Complex Needs of the Health Care Team: Identification of Nurse-Team Communication Practices Perceived to Enhance Patient Outcomes. Qualitative Health Research. 2010;20(1):15–28. doi: 10.1177/1049732309355289. PubMed PMID: WOS:000272852200003.

21. Cranstoun D, Baliousis M, Merdian HL, Rennoldson M. Nurse-Led Psychological Interventions For Depression In Adult Cancer Patients: A Systematic Review And MetaAnalysis of Randomized Controlled Trials. Journal of Pain and Symptom Management. 2024;68(1):e21–e35. doi: 10.1016/j.jpainsymman.2024.03.028. PubMed PMID: WOS:001252452100001.

22. Mohammadi F, Hosseiny SMM, Khazaei S, Esfahani H, Soltani K, Masumi SM. Effect of nurse-led intervention programs based on spiritual care on the resilience and death anxiety in parents of children and adolescents with cancer, a randomized clinical trial. Bmc Psychology. 2025;13(1). doi: 10.1186/s40359-025-03179-w. PubMed PMID: WOS:001546443900003.

23. Wu ZL, Li XL, Huang YX, Huang KL, Xiao B, Chi YF, et al. Effects of a Nurse-Led Cognitive Behavioral Intervention for Parents of Children With Epilepsy. Pediatric Neurology. 2024;154:70–8. doi: 10.1016/j.pediatrneurol.2024.03.003. PubMed PMID: WOS:001221735000001.

24. Soleimani MA, Alimoradi Z, Kazemi S. The impact of dignity therapy on demoralization syndrome in patients undergoing chemotherapy: A randomized controlled trial exploring a nurse-led psychotherapeutic approach in oncology care. European Journal of Oncology Nursing. 2025;77. doi: 10.1016/j.ejon.2025.102934. PubMed PMID: WOS:001541524200001.

25. Hróbjartsson A, Boutron I, Hopewell S, Moher D, Schulz KF, Collins GS, et al. SPIRIT 2025 explanation and elaboration: updated guideline for protocols of randomised trials. BMJ. 2025;389:e081660. doi: 10.1136/bmj-2024-081660.

26. Peng Y. The Cultural Adaptation and Initial Application ofSurvivorship Cancer Competently Intervention Programfor Families of Children with [Master’s thesis]: Central south university; 2024.

27. Steliarova-Foucher E, Stiller C, Lacour B, Kaatsch P. International classification of childhood cancer. Cancer. 2005;103(7):1457–67.

28. Steinberg AM, Brymer MJ, Decker KB, Pynoos RS. The University of California at Los Angeles post-traumatic stress disorder reaction index. Current psychiatry reports. 2004;6(2):96–100.

29. Ferrafiat V, Soleimani M, Chaumette B, Martinez A, Guilé J-M, Keeshin B, et al. Use of prazosin for pediatric post-traumatic stress disorder with nightmares and/or sleep disorder: case series of 18 patients prospectively assessed. Frontiers in Psychiatry. 2020;11:724.

30. Lin F, Jin C, Suman W, Zhengkui L. Validity and reliability of the Chinese Version of the University of California at Los Angeles Posttraumatic Stress Disorder Reaction Index for DSM-IV (Revision 1, Children version) Chinese Mental Health Journal/Zhongguo Xinli Weisheng Zazhi. 2018;32(2).

31. Weiss DS. The impact of event scale: revised. Cross-cultural assessment of psychological trauma and PTSD: Springer; 2007. p. 219–38.

32. Creamer M, Bell R, Failla S. Psychometric properties of the impact of event scale—revised. Behaviour research and therapy. 2003;41(12):1489–96.

33. Meyerson DA, Grant KE, Carter JS, Kilmer RP. Posttraumatic growth among children and adolescents: A systematic review. Clinical psychology review. 2011;31(6):949–64.

34. Schepers SA, Okado Y, Russell K, Long AM, Phipps S. Adjustment in childhood cancer survivors, healthy peers, and their parents: The mediating role of the parent–child relationship. Journal of pediatric psychology. 2019;44(2):186–96.

35. Wang J, Chen Y, Wang Y, Liu X. Revision of the posttraumatic growth inventory and testing its reliability and validity. J Nurs Sci. 2011;26(14):26–8.

36. Wells SY, Morland LA, Torres EM, Kloezeman K, Mackintosh M-A, Aarons GA. The development of a brief version of the Posttraumatic Cognitions Inventory (PTCI-9). Assessment. 2019;26(2):193–208.

37. Foa EB, Ehlers A, Clark DM, Tolin DF, Orsillo SM. The posttraumatic cognitions inventory (PTCI): Development and validation. Psychological assessment. 1999;11(3):303.

38. Zhan N, Gao C, Cao Y, Li F, Geng F. Factor structure, measurement invariance, and psychometric properties of the Posttraumatic Cognitions Inventory (PTCI) and its brief version (PTCI-9) in Chinese adolescents and adults. Psychological Assessment. 2024;36(4):291.

39. Spitzer RL, Kroenke K, Williams JB, Löwe B. A brief measure for assessing generalized anxiety disorder: the GAD-7. Archives of internal medicine. 2006;166(10):1092–7.

40. Kroenke K, Spitzer RL, Williams JB. The PHQ-9: validity of a brief depression severity measure. Journal of general internal medicine. 2001;16(9):606–13.

41. Knafl K, Deatrick JA, Gallo A, Dixon J, Grey M, Knafl G, et al. Assessment of the psychometric properties of the family management measure. Journal of pediatric psychology. 2011;36(5):494–505.

42. McCubbin HI, Thompson AI, McCubbin MA. Family assessment: Resiliency, coping and adaptation: Inventories for research and practice. (No Title). 1996.

43. Liu Y, Yang J, Ye B, Shen Q, Zhu J, Chen M. Reliability and validity of the Chinese version of family hardiness index. J Nurs Adm. 2014;11(14):770–2.

44. Varni JW, Burwinkle TM, Katz ER, Meeske K, Dickinson P. The PedsQL™ in pediatric cancer: reliability and validity of the pediatric quality of life inventory™ generic core scales, multidimensional fatigue scale, and cancer module. Cancer. 2002;94(7):2090–106.

45. Ji Y, Chen S, Li K, Xiao N, Yang X, Zheng S, et al. Measuring health-related quality of life in children with cancer living in Mainland China: feasibility, reliability and validity of the Chinese Mandarin version of PedsQL 4.0 Generic Core Scales and 3.0 Cancer Module. Health and quality of life outcomes. 2011;9(1):103.

46. Buysse DJ, Reynolds CF, 3rd, Monk TH, Berman SR, Kupfer DJ. The Pittsburgh Sleep Quality Index: a new instrument for psychiatric practice and research. Psychiatry Res. 1989;28(2):193–213. doi: 10.1016/0165-1781(89)90047-4. PubMed PMID: 2748771.

47. Ho KY, Lam KKW, Xia W, Chung JOK, Cheung AT, Ho LLK, et al. Psychometric properties of the Chinese version of the Pittsburgh Sleep Quality Index (PSQI) among Hong Kong Chinese childhood cancer survivors. Health and Quality of Life Outcomes. 2021;19(1). doi: 10.1186/s12955-021-01803-y. PubMed PMID: WOS:000670272500001.

48. Fan B, Meng L, Bai L, Du Y, Ding H, Chen Y, et al. Prolonged periods of shallow sleep are associated with diabetic carotid atherosclerosis. European Journal of Medical Research. 2025;30(1):647. doi: 10.1186/s40001-025-02923-7.

49. Harrison SL, Buckley BJR, Zheng Y, Hill A, Hlaing T, Davies R, et al. Evaluation of Huawei smart wearables for detection of atrial fibrillation in patients following ischemic stroke: The Liverpool-Huawei stroke study. American Heart Journal. 2023;257:103–10. doi: 10.1016/j.ahj.2022.12.004.

50. Glasgow RE, Vogt TM, Boles SM. Evaluating the public health impact of health promotion interventions: the RE-AIM framework. Am J Public Health. 1999;89(9):1322–7. doi: 10.2105/ajph.89.9.1322. PubMed PMID: 10474547; PubMed Central PMCID: PMCPMC1508772.

51. Andriessen C, Blom MT, van Hoek BACE, de Boer AW, Denig P, de Wit GA, et al. A deprescribing programme aimed to optimise blood glucose-lowering medication in older people with type 2 diabetes mellitus, the OMED2-study: the study protocol for a randomised controlled trial. Trials. 2024;25(1):505. doi: 10.1186/s13063-024-08249-9.

52. Rørbech JT, Dreyer P, Enskär K, Haslund-Thomsen H, Jensen CS. Nursing interventions for pediatric patients with cancer and their families: A scoping review. International Journal of Nursing Studies. 2024;160:104891. doi: 10.1016/j.ijnurstu.2024.104891.

53. Osborn T. The psychosocial impact of parental cancer on children and adolescents: a systematic review. Psycho-Oncology. 2007;16(2):101–26. doi: 10.1002/pon.1113. PubMed PMID: WOS:000244607400003.

54. Bidstrup PE, Salem H, Andersen EW, Schmiegelow K, Rosthøj S, Wehner PS, et al. Effects on Pediatric Cancer Survivors: The FAMily-Oriented Support (FAMOS) Randomized Controlled Trial. Journal of Pediatric Psychology. 2022;48(1):29–38. doi: 10.1093/jpepsy/jsac062.

55. Zhang Y, Zhang Z, Peng Y, Zhang W, Ma G, Lin S, et al. Impact of technology- and parent-based psychosocial interventions on family dynamics factors in children with cancer: A systematic review. PLOS ONE. 2025;20(5):e0323483. doi: 10.1371/journal.pone.0323483.

